# Associations of diarised sleep onset time, period and duration with total and central adiposity in a biethnic sample of young children: the Born in Bradford study

**DOI:** 10.1101/2020.09.30.20193888

**Authors:** Paul J Collings, Jane E Blackwell, Elizabeth Pal, Helen L Ball, John Wright

## Abstract

**Objectives:** To investigate associations of sleep timing, period and duration with total and abdominal adiposity in a biethnic sample of children aged 18 and 36 months (m)

**Design:** Cross-sectional observational study

**Setting:** The Born in Bradford 1000 study, UK

**Participants:** Children aged approximately 18m (*n*=209; 40.2% South Asian; 59.8% White) and 36m (*n*=162; 40.7% South Asian; 59.3% White)

**Primary and secondary outcome measures:** Parents completed a 3-day sleep diary from which children’s average daily sleep onset time, period and duration were calculated. Weekday to weekend differences in sleep parameters were also derived. As outcomes, indices of total (BMI z-score and sum of 2-skinfolds) and abdominal adiposity (waist circumference) were measured. Adjusted regression was used to quantify associations of sleep parameters with adiposity by age group and ethnicity.

**Results:** The average daily sleep onset time was markedly later in South Asian (9:26pm ± 68 mins) than White children (7:41pm ± 48 mins). Later sleep onset was associated with lower BMI z-score and sum of 2-skinfolds in White children aged 18m. In contrast, later sleep onset was associated with higher BMI z-score in South Asian children aged 36m. For weekday to weekend differences, longer sleep duration and later sleep onset on weekends than weekdays were both associated with higher total and abdominal adiposity in South Asian children aged 18m. On the contrary, compared to consistent sleep onset times, going to sleep ≥20 minutes later on weekends than weekdays was associated with lower waist circumference in White children aged 18m.

**Conclusions:** Sleep timing is associated with total and central adiposity in young children but associations differ by age group and ethnicity. Sleep onset times and regular sleep schedules may be important for obesity prevention.

## Introduction

Many studies have investigated associations of sleep duration or the sleep period (here defined as elapsed time between sleep onset and termination regardless of waking intervals) with adiposity in school-aged children and youth [1,2]. Few such studies have been conducted in infants and toddlers despite sleep problems and sleep loss being prevalent in children aged <36 months (m) [3]. Beyond sleep duration, it is beginning to be recognised that sleep is a multidimensional construct of partly overlapping dimensions, and that parameters such as sleep timing may also be influential with regard to adiposity level and obesity risk [4].

Aspects of sleep timing include bedtimes, time of sleep onset, and variability in sleeping pattern such as differences between weekday and weekend sleep schedules. Preliminary evidence indicates that later bedtimes are associated with higher weight status in pre-schoolers [5–8] and primary school-aged children [9], and that later sleep timing on weekends than weekdays is correlated with higher risk of overweight and obesity [10]. Hypothesised mechanisms include circadian dysrhythmia causing hormone-mediated preferences for unhealthy calorific foodstuffs [11,12], the effect of which is compounded by later sleep onset enabling more time to eat, more opportunity for screen-based sedentary time such as TV viewing [13], and lower daily physical activity and higher sedentariness due to delayed waking and fatigue [14]. More studies need to investigate the independent associations of myriad sleep parameters with childhood adiposity and to ascertain the underlying mechanisms of action. A research focus on young children is important, to determine when associations between sleep parameters with adiposity first emerge, and to provide information about how to intervene by identifying modifiable target behaviours [4].

Early childhood is a critical window for obesity prevention because it is a period when rapid weight gain occurs and behavioural patterns are first established [15,16]. We are aware of only one study that has investigated associations of sleep duration and bedtimes with adiposity in early childhood. That study was conducted in children from America, Australia and New Zealand and the vast majority of mothers were of White ethnicity [17]. Investigation of ethnic minority and socioeconomically impoverished populations is needed. Children of non-White ethnicity and children from deprived backgrounds have unfavourable sleeping patterns and are high-risk for early onset obesity [18,19]. We have previously shown that UK South Asian children who live in a deprived urban setting go to bed much later than White children from the same area [20]. It is important to determine if shifted sleep schedules may in part explain why obesity rates are higher in South Asian than White children in the UK [21], and why South Asian school-aged children have more centrally-stored adiposity which is metabolically harmful [22]. This information could help reduce ethnic disparities in disease risk by facilitating behaviour change interventions and policy guidelines that are tailored to the needs of specific ethnic groups.

This study examined independent associations of a broad range of sleep parameters including diarised sleep onset time, period and duration, and weekday to weekend variations in sleep parameters, with total and central adiposity in a young biethnic sample of UK children from a deprived urban setting.

## Methods

This study was conducted in Bradford which is the fifth largest local authority in England and one of the most deprived and ethnically diverse cities in the UK [23]. Bradford has an overweight and obesity prevalence of 21.8% in 4 to 5 year old children and 38.4% in 10 to 11 year olds [24]. The Born in Bradford (BiB) 1000 study [25], nested within the larger BiB pregnancy cohort [26], aims to investigate modifiable risk factors for childhood obesity. Pregnant women (*n*=1916) were invited to the BiB 1000 study when they attended routine hospital appointments, 90.6% accepted the invitation (*n*=1735). Consent to medical records access was provided and periodic postnatal assessments were carried out when the women’s offspring were approximately 6, 12, 18, 24, and 36m old. Parents who participated when children were aged about 18m (*n*=1228; 70.7% of all BiB 1000 participants) or 36m (*n*=1232; 71%) were asked to complete a sleep diary for their child. At each timepoint diaries were completed for approximately 15% of all BiB 1000 participants (18m: *n*=276; 36m: *n*=262). The diary data were collected between October 2010 and September 2012. This complete-case analysis included only children with sleep diary data, concurrent adiposity measurements and information about potentially important covariates. Maternal ethnicity was used as a proxy for child ethnicity, and children with a mother belonging to an ethnic group other than South Asian (Pakistani / Indian / Bangladeshi) or White (British / other) were excluded due to small numbers. All children were born in the UK, most children who were of South Asian heritage had Pakistani origin mothers (85%), and most children who were White had White British mothers (94%). For brevity we refer to children as being of South Asian or White ethnicity. The final sample included 209 children aged 18m (12% of all BiB 1000 participants) and 162 children aged 36m (9.3%). Ethical approval for all aspects of the study was granted by Bradford Research Ethics Committee (Reference 07/H1302/112) and all mothers provided informed written consent for participation.

### Sleep parameters

Parents completed sleep diaries, providing free-text responses about the time their child fell asleep on an evening and woke-up the next morning. The sleep period was calculated as elapsed time between sleep onset and waking the next day. Because the sleep period does not account for periods of overnight waking which can be common in young children [27] parents also reported their child’s overnight sleep duration. Parents completed diaries on two weekdays and a weekend day. Mean weekday values were calculated by averaging data for the two weekdays; thereafter daily averages over the course of a week were calculated using weekday-weekend weighting (in the ratio of 5:2). Weekday to weekend differences in sleep parameters were calculated by subtraction (weekend – weekday values).

### Adiposity markers

Child weight and height were assessed by trained researchers and BMI (kg/m^2^) and *z*-scores were calculated [28]. Waist circumference was measured at the level of the navel. Skinfolds of the left triceps and subscapular were measured to the nearest 0.1mm and summed. Reliability metrics indicated good intra- and inter-observer technical error for measurements [29].

### Covariates

Potential confounders and mediators of associations were selected based on previous evidence linking them with sleep and obesity [15,18]. A multidimensional marker of socioeconomic status was synthesised from information collated during interviews with parents (usually mothers) about their education, employment, housing tenure, financial situation, and ownership of goods. For the purposes of this study, from an initial five categories [30], children were classified as belonging to one of three parental socioeconomic status groups: least deprived (least socioeconomically deprived and most educated), moderately deprived (employed and not materially deprived / employed but no access to money) and most deprived (benefits but not materially deprived / most economically deprived). Maternal age was self-reported in pregnancy. The number of previous births (used to group mothers as primaparous (pregnant for the first time) or not), child gender, gestational age, and birth weight were extracted from medical records. Maternal height and weight were measured at the 18 and 36m timepoints and were used to calculate maternal BMI. Parents reported whether or not their child napped on sleep diary reporting days, these data were used to create three categories of napping frequency (never: napped zero days; occasionally: napped 1 or 2 days; every day: napped on all three days). On each sleep diary reporting day, parents also provided free-text responses about the time their child ate their last meal of the day. Mean weekday values were calculated by averaging data across the two weekdays, and weekday-weekend weighting was used to calculate the average daily time that the child ate their last meal; weekday to weekend differences were calculated by subtraction. Infant dietary data were collected using a validated food frequency questionnaire that was modified to include ethnic-specific foodstuffs; dietary constructs indicative of unhealthy snacking (frequency of biscuit, crisps, cakes, sweets, chocolate, sugar-sweetened beverage consumption) and fruit and vegetable consumption in the previous 4-12 weeks were derived [20]. Parents reported the number of hours their child watched TV on a typical weekday and weekend day; weekday-weekend weighting was used to calculate average daily TV viewing and weekday to weekend differences were calculated [31]. When children were aged 36m a parent-reported Early Years Physical Activity Questionnaire (EY-PAQ) was used to estimate children’s physical activity level [32]. Calendar dates of the 18 and 36m assessments were recorded and categorised by season (summer / autumn or spring / winter).

### Patient and public involvement (PPI)

The BiB research team regularly convenes a Parent Governors PPI group, BiB participants now aged 10 to 12 years can become Young Ambassadors, and BiB runs regular community events and science festivals (https://borninbradford.nhs.uk/news-events/events/). These committees and events help to shape our research by allowing us to learn about the opinions, concerns and ideas of the community. A study author (JEB) also recently chaired a PPI event held in conjunction with The Sleep Charity [33]. Attendees were parents of children with sleep difficulties, staff who work in residential settings with children, sleep practitioners and representatives from parent and carer forums. Emerging themes from the session included: 1) there is a lack of information about assisting development of healthy sleep routines in infants, 2) more information and knowledge about the consequences of poor sleep in infants is needed, 3) early intervention is vital and parents of young children should be offered evidence-based sleep interventions for their child.

### Statistics

Correlations between individual sleep parameters and adiposity markers were calculated using Pearson or Spearman methods as appropriate [34]. Adjusted linear regression was used to investigate cross-sectional associations between sleep parameters with adiposity. Model 1 adjusted associations for sex, age, socioeconomic status, maternal pregnancy age, parity, gestational age, birthweight, maternal BMI, season of measurement, and napping frequency. Model 2 further adjusted for potential confounding or mediating factors, including unhealthy snacking, fruit and vegetable intake, and TV viewing. Model 3 mutually adjusted sleep period and duration variables for sleep onset time, and vice versa. All analyses were performed stratified by ethnic group and age group. Interaction effects between sleep period and duration with sleep onset time were examined by introducing multiplicative terms to models (eg. sleep onset*duration). Nonlinearity of associations was examined by adding quadratic terms for sleep parameters. There was some of evidence for nonlinear associations between average daily sleep onset and weekday to weekend differences in sleep onset with waist circumference and sum of 2-skinfolds, hence sleep onset variables were trichotomised and adjusted linear regression was used to estimate group marginal means (and 95% confidence intervals (CI)) for these outcomes. Data for the sum of 2-skinfolds were slightly skewed, but because results were consistent regardless of whether the data were log-transformed or not, results based on the untransformed data are presented. Sensitivity analyses further adjusted associations for the time of last meal and physical activity (not applicable to 18m models when physical activity was not assessed), potentially important covariates but missing data; models with waist circumference or sum of 2-skinfolds as dependent variables were further adjusted for height. Analyses were performed with Stata/SE 16.1 software (StataCorp, College Station, TX); *a priori p*<0.05 was deemed statistically significant but the results are interpreted with emphasis on the range of plausible values of associations as shown by confidence intervals [35].

## Results

### Sample characteristics

Participant details are shown in **Table 1**. Child ages ranged from 16.4 to 22.8m and 35.5 to 39.5m at each respective timepoint. The average daily sleep onset time was consistently after 9pm in South Asian children and before 8pm in White children. Sleep onset time was inversely correlated with the sleep period (South Asian children aged 18m: −0.52; South Asian children aged 36m: −0.39; White children aged 18m: −0.52; White children aged 36m: - 0.37) and to a lesser extent sleep duration (South Asian children aged 18m: −0.41; South Asian children aged 36m: −0.10; White children aged 18m: −0.38; White children aged 36m: - 0.27). Correlations between the sleep period and sleep duration were weaker in South Asian than White children (South Asian children aged 18m: 0.72; South Asian children aged 36m: 0.66; White children aged 18m: 0.81; White children aged 36m: 0.83). Correlations between adiposity markers differed by ethnic group and age, but consistently BMI z-score was more strongly correlated with waist circumference (South Asian children aged 18m: 0.69; South Asian children aged 36m: 0.71; White children aged 18m: 0.61; White children aged 36m: 0.79) than the sum of 2-skinfolds (South Asian children aged 18m: 0.48; South Asian children aged 36m: 0.28; White children aged 18m: 0.44; White children aged 36m: 0.53); correlations between waist circumference and the sum of 2-skinfolds were weakest (South Asian children aged 18m: 0.42; South Asian children aged 36m: 0.19; White children aged 18m: 0.26; White children aged 36m: 0.47).

**Table 1.**
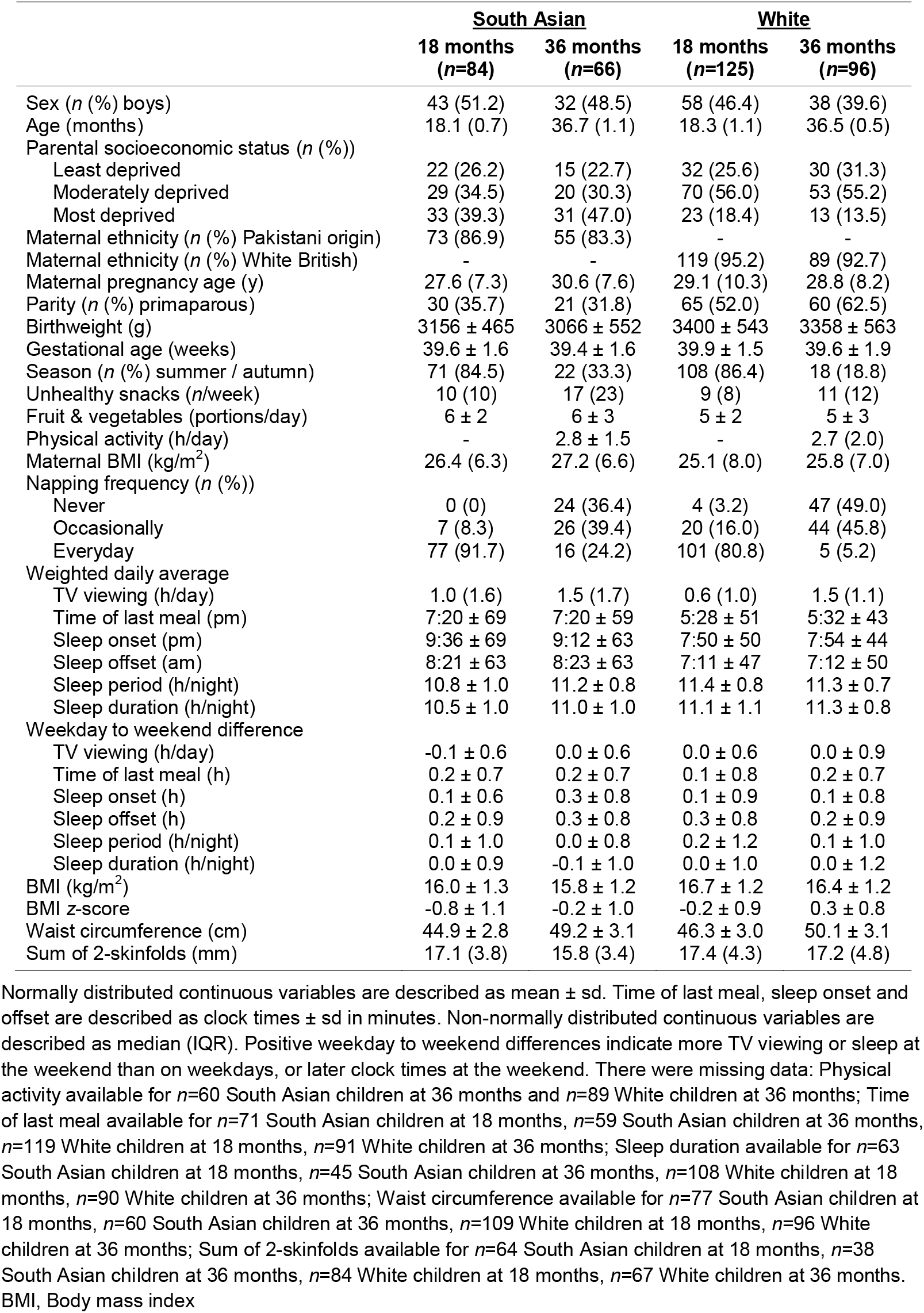
Description of study participants.

|  | **South Asian** |  | **White** |  |
| --- | --- | --- | --- | --- |
|  | **18 months**<br>**(n=84)** | **36 months**<br>**(n=66)** | **18 months**<br>**(n=125)** | **36 months**<br>**(n=96)** |
| Sex (*n* (%) boys) | 43 (51.2) | 32 (48.5) | 58 (46.4) | 38 (39.6) |
| Age (months) | 18.1 (0.7) | 36.7 (1.1) | 18.3 (1.1) | 36.5 (0.5) |
| Parental socioeconomic status (*n* (%)) |  |  |  |  |
| Least deprived | 22 (26.2) | 15 (22.7) | 32 (25.6) | 30 (31.3) |
| Moderately deprived | 29 (34.5) | 20 (30.3) | 70 (56.0) | 53 (55.2) |
| Most deprived | 33 (39.3) | 31 (47.0) | 23 (18.4) | 13 (13.5) |
| Maternal ethnicity (*n* (%) Pakistani origin) | 73 (86.9) | 55 (83.3) | - | - |
| Maternal ethnicity (*n* (%) White British) | - | - | 119 (95.2) | 89 (92.7) |
| Maternal pregnancy age (y) | 27.6 (7.3) | 30.6 (7.6) | 29.1 (10.3) | 28.8 (8.2) |
| Parity (*n* (%) primiparous) | 30 (35.7) | 21 (31.8) | 65 (52.0) | 60 (62.5) |
| Birthweight (g) | 3156 ± 465 | 3066 ± 552 | 3400 ± 543 | 3358 ± 563 |
| Gestational age (weeks) | 39.6 ± 1.6 | 39.4 ± 1.6 | 39.9 ± 1.5 | 39.6 ± 1.9 |
| Season (*n* (%) summer / autumn) | 71 (84.5) | 22 (33.3) | 108 (86.4) | 18 (18.8) |
| Unhealthy snacks (*n*/week) | 10 (10) | 17 (23) | 9 (8) | 11 (12) |
| Fruit & vegetables (portions/day) | 6 ± 2 | 6 ± 3 | 5 ± 2 | 5 ± 3 |
| Physical activity (h/day) | - | 2.8 ± 1.5 | - | 2.7 (2.0) |
| Maternal BMI (kg/m<sup>2</sup>) | 26.4 (6.3) | 27.2 (6.6) | 25.1 (8.0) | 25.8 (7.0) |
| Napping frequency (*n* (%)) |  |  |  |  |
| Never | 0 (0) | 24 (36.4) | 4 (3.2) | 47 (49.0) |
| Occasionally | 7 (8.3) | 26 (39.4) | 20 (16.0) | 44 (45.8) |
| Everyday | 77 (91.7) | 16 (24.2) | 101 (80.8) | 5 (5.2) |
| Weighted daily average |  |  |  |  |
| TV viewing (h/day) | 1.0 (1.6) | 1.5 (1.7) | 0.6 (1.0) | 1.5 (1.1) |
| Time of last meal (pm) | 7:20 ± 69 | 7:20 ± 59 | 5:28 ± 51 | 5:32 ± 43 |
| Sleep onset (pm) | 9:36 ± 69 | 9:12 ± 63 | 7:50 ± 50 | 7:54 ± 44 |
| Sleep offset (am) | 8:21 ± 63 | 8:23 ± 63 | 7:11 ± 47 | 7:12 ± 50 |
| Sleep period (h/night) | 10.8 ± 1.0 | 11.2 ± 0.8 | 11.4 ± 0.8 | 11.3 ± 0.7 |
| Sleep duration (h/night) | 10.5 ± 1.0 | 11.0 ± 1.0 | 11.1 ± 1.1 | 11.3 ± 0.8 |
| Weekday to weekend difference |  |  |  |  |
| TV viewing (h/day) | -0.1 ± 0.6 | 0.0 ± 0.6 | 0.0 ± 0.6 | 0.0 ± 0.9 |
| Time of last meal (h) | 0.2 ± 0.7 | 0.2 ± 0.7 | 0.1 ± 0.8 | 0.2 ± 0.7 |
| Sleep onset (h) | 0.1 ± 0.6 | 0.3 ± 0.8 | 0.1 ± 0.9 | 0.1 ± 0.8 |
| Sleep offset (h) | 0.2 ± 0.9 | 0.3 ± 0.8 | 0.3 ± 0.8 | 0.2 ± 0.9 |
| Sleep period (h/night) | 0.1 ± 1.0 | 0.0 ± 0.8 | 0.2 ± 1.2 | 0.1 ± 1.0 |
| Sleep duration (h/night) | 0.0 ± 0.9 | -0.1 ± 1.0 | 0.0 ± 1.0 | 0.0 ± 1.2 |
| BMI (kg/m<sup>2</sup>) | 16.0 ± 1.3 | 15.8 ± 1.2 | 16.7 ± 1.2 | 16.4 ± 1.2 |
| BMI z-score | -0.8 ± 1.1 | -0.2 ± 1.0 | -0.2 ± 0.9 | 0.3 ± 0.8 |
| Waist circumference (cm) | 44.9 ± 2.8 | 49.2 ± 3.1 | 46.3 ± 3.0 | 50.1 ± 3.1 |
| Sum of 2-skinfolds (mm) | 17.1 (3.8) | 15.8 (3.4) | 17.4 (4.3) | 17.2 (4.8) |

Normally distributed continuous variables are described as mean ± sd. Time of last meal, sleep onset and offset are described as clock times ± sd in minutes. Non-normally distributed continuous variables are described as median (IQR). Positive weekday to weekend differences indicate more TV viewing or sleep at the weekend than on weekdays, or later clock times at the weekend. There were missing data: Physical activity available for *n*=60 South Asian children at 36 months and *n*=89 White children at 36 months; Time of last meal available for *n*=71 South Asian children at 18 months, *n*=59 South Asian children at 36 months, *n*=119 White children at 18 months, *n*=91 White children at 36 months; Sleep duration available for *n*=63 South Asian children at 18 months, *n*=45 South Asian children at 36 months, *n*=108 White children at 18 months, *n*=90 White children at 36 months; Waist circumference available for *n*=77 South Asian children at 18 months, *n*=60 South Asian children at 36 months, *n*=109 White children at 18 months, *n*=96 White children at 36 months; Sum of 2-skinfolds available for *n*=64 South Asian children at 18 months, *n*=38 South Asian children at 36 months, *n*=84 White children at 18 months, *n*=67 White children at 36 months. BMI, Body mass index

### South Asian children

**Table 2** shows that in South Asian children aged 18m there were no significant associations between average daily sleep parameters with adiposity. In children aged 36m, there was some evidence in model 3 that later daily sleep onset was associated with higher BMI *z*-score; the association attenuated when further adjusted for final mealtime (β=0.2 (95% CI:-0.2 to 0.6), *p*=0.34). There were no other significant associations for average daily sleep parameters. **Figure 1** presents the group marginal means for waist circumference and the sum of 2-skinfolds stratified by sleep onset categories; there were no group differences. With regard to weekday to weekend differences, Table 2 shows that longer sleep duration on weekends than weekdays was consistently associated with higher adiposity in South Asian children aged 18m, and in the same group of children, later sleep onset on weekends than weekdays was associated with larger waist circumference and sum of 2-skinfolds. In children aged 36m, later sleep onset on weekends than weekdays was associated with larger sum of 2-skinfolds in model 2, but the association attenuated when further adjusted for the sleep period. **Figure 2** shows the adjusted marginal means for waist circumference and sum of 2-skinfolds stratified by three categories of weekday to weekend differences in sleep onset. There was some evidence that compared to children with consistent sleep onset times (±20 minutes of each other), children aged 18m who went to sleep ≥20 minutes later on weekends than weekdays had larger waist circumferences (1.5 (−0.1 to 3.0) cm, *p*=0.060) and sum of 2-skinfolds (1.9 (−0.2 to 4.1) mm, *p*=0.078). Similarly, there was some indication that aged 36m, children who went to sleep ≥20 minutes later on weekends than weekdays had larger sum of 2-skinfolds (2.0 (−0.2 to 4.3) mm, *p*=0.074).

**Table 2.** Associations of sleep onset, period and duration with adiposity in South Asian children, stratified by age group.

|  |  | Body mass index z-score |  |  |  |  | Waist circumference (cm) |  |  |  |  | Sum of 2-skinfolds (mm) |  |  |  |  |
| --- | --- | --- | --- | --- | --- | --- | --- | --- | --- | --- | --- | --- | --- | --- | --- | --- |
|  |  | Weighted daily |  | average | Weekday to weekend difference |  | Weighted daily |  | average | Weekday to weekend difference |  | Weighted daily |  | average | Weekday to weekend difference |  |
| | | *n* | $\beta$ (95% CI) | *p* | $\beta$ (95% CI) | *p* | *n* | $\beta$ (95% CI) | *p* | $\beta$ (95% CI) | *p* | *n* | $\beta$ (95% CI) | *p* | $\beta$ (95% CI) | *p* |
| 18 months | Model 1 |  |  |  |  |  |  |  |  |  |  |  |  |  |  |  |
|  | Sleep onset | 84 | -0.1 (-0.3 to 0.2) | 0.66 | 0.1 (-0.4 to 0.5) | 0.76 | 77 | -0.4 (-1.0 to 0.2) | 0.24 | 0.8 (-0.3 to 1.8) | 0.17 | 64 | -0.2 (-1.1 to 0.7) | 0.68 | 1.3 (-0.1 to 2.8) | 0.070 |
|  | Sleep period | 84 | 0.2 (-0.1 to 0.4) | 0.16 | 0.0 (-0.3 to 0.3) | 0.84 | 77 | 0.3 (-0.3 to 1.0) | 0.30 | 0.0 (-0.7 to 0.8) | 0.91 | 64 | 0.2 (-0.7 to 1.0) | 0.70 | -0.3 (-1.4 to 0.8) | 0.56 |
|  | Sleep duration | 63 | 0.2 (-0.2 to 0.5) | 0.29 | 0.3 (-0.1 to 0.6) | 0.17 | 58 | 0.3 (-0.6 to 1.2) | 0.54 | 0.6 (-0.4 to 1.7) | 0.21 | 49 | 0.5 (-0.7 to 1.6) | 0.41 | 0.8 (-0.6 to 2.2) | 0.26 |
|  | Model 2 |  |  |  |  |  |  |  |  |  |  |  |  |  |  |  |
|  | Sleep onset | 84 | 0.0 (-0.2 to 0.3) | 0.93 | 0.0 (-0.4 to 0.4) | 0.93 | 77 | -0.3 (-0.9 to 0.4) | 0.41 | 0.7 (-0.4 to 1.8) | 0.21 | 64 | -0.1 (-0.9 to 0.8) | 0.90 | **1.6 (0.1 to 3.1)** | **0.036** |
|  | Sleep period | 84 | 0.2 (-0.1 to 0.4) | 0.22 | 0.1 (-0.2 to 0.4) | 0.59 | 77 | 0.3 (-0.4 to 1.0) | 0.39 | 0.3 (-0.5 to 1.1) | 0.47 | 64 | 0.1 (-0.8 to 1.0) | 0.89 | -0.2 (-1.4 to 0.9) | 0.72 |
|  | Sleep duration | 63 | 0.2 (-0.1 to 0.5) | 0.27 | 0.3 (0.0 to 0.7) | 0.066 | 58 | 0.3 (-0.6 to 1.2) | 0.55 | 0.9 (-0.1 to 1.9) | 0.075 | 49 | 0.3 (-0.9 to 1.5) | 0.61 | 0.9 (-0.6 to 2.3) | 0.23 |
|  | Model 3 |  |  |  |  |  |  |  |  |  |  |  |  |  |  |  |
|  | Sleep onset<sup>1</sup> | 84 | 0.1 (-0.2 to 0.4) | 0.40 | 0.0 (-0.5 to 0.5) | 0.99 | 77 | -0.2 (-0.9 to 0.6) | 0.65 | 1.2 (-0.1 to 2.4) | 0.076 | 64 | 0.0 (-1.1 to 1.1) | 0.96 | **1.8 (0.1 to 3.5)** | **0.034** |
|  | Sleep onset<sup>2</sup> | 63 | 0.2 (-0.1 to 0.5) | 0.23 | 0.4 (-0.1 to 0.8) | 0.12 | 58 | 0.2 (-0.6 to 1.1) | 0.58 | **1.8 (0.6 to 2.9)** | **0.004** | 49 | 0.0 (-0.9 to 1.1) | 0.92 | **1.7 (0.3 to 3.1)** | **0.022** |
|  | Sleep period | 84 | 0.2 (-0.1 to 0.5) | 0.14 | 0.1 (-0.2 to 0.4) | 0.63 | 77 | 0.2 (-0.6 to 1.0) | 0.60 | 0.7 (-0.2 to 1.5) | 0.14 | 64 | 0.0 (-1.1 to 1.1) | 0.93 | 0.3 (-0.9 to 1.5) | 0.61 |
|  | Sleep duration | 63 | 0.3 (-0.1 to 0.6) | 0.14 | **0.4 (0.1 to 0.8)** | **0.024** | 58 | 0.3 (-0.6 to 1.3) | 0.46 | **1.2 (0.3 to 2.2)** | **0.012** | 49 | 0.3 (-0.9 to 1.5) | 0.61 | 1.4 (0.0 to 2.9) | 0.056 |
| 36 months | Model 1 |  |  |  |  |  |  |  |  |  |  |  |  |  |  |  |
|  | Sleep onset | 66 | 0.1 (-0.1 to 0.4) | 0.17 | -0.3 (-0.6 to 0.0) | 0.085 | 60 | -0.1 (-1.0 to 0.7) | 0.74 | **-1.2 (-2.4 to 0.0)** | **0.048** | 38 | -0.3 (-1.3 to 0.7) | 0.52 | 0.9 (-0.3 to 2.1) | 0.14 |
|  | Sleep period | 66 | 0.1 (-0.2 to 0.4) | 0.50 | 0.2 (-0.1 to 0.5) | 0.14 | 60 | -0.4 (-1.7 to 0.8) | 0.48 | **1.1 (0.0 to 2.3)** | **0.047** | 38 | 0.0 (-1.2 to 1.3) | 0.96 | -0.3 (-1.8 to 1.1) | 0.64 |
|  | Sleep duration | 45 | 0.2 (-0.1 to 0.5) | 0.15 | 0.0 (-0.4 to 0.3) | 0.82 | 42 | 0.5 (-0.6 to 1.5) | 0.35 | 0.3 (-1.0 to 1.6) | 0.63 | 26 | 0.2 (-0.9 to 1.2) | 0.71 | -0.1 (-1.8 to 1.6) | 0.92 |
|  | Model 2 |  |  |  |  |  |  |  |  |  |  |  |  |  |  |  |
|  | Sleep onset | 66 | 0.2 (-0.1 to 0.4) | 0.13 | -0.2 (-0.6 to 0.1) | 0.15 | 60 | 0.1 (-0.8 to 1.0) | 0.86 | -1.0 (-2.2 to 0.2) | 0.11 | 38 | -0.2 (-1.4 to 1.0) | 0.75 | **1.3 (0.1 to 2.5)** | **0.033** |
|  | Sleep period | 66 | 0.1 (-0.2 to 0.5) | 0.42 | 0.2 (-0.1 to 0.5) | 0.14 | 60 | -0.4 (-1.7 to 0.9) | 0.55 | **1.2 (0.1 to 2.3)** | **0.032** | 38 | 0.1 (-1.2 to 1.4) | 0.84 | -1.1 (-2.7 to 0.4) | 0.15 |
|  | Sleep duration | 45 | 0.2 (-0.1 to 0.5) | 0.27 | 0.1 (-0.3 to 0.5) | 0.67 | 42 | 0.3 (-0.8 to 1.4) | 0.55 | 0.5 (-0.8 to 1.9) | 0.44 | 26 | 0.0 (-1.1 to 1.0) | 0.97 | -0.8 (-3.1 to 1.5) | 0.43 |
|  | Model 3 |  |  |  |  |  |  |  |  |  |  |  |  |  |  |  |
|  | Sleep onset<sup>1</sup> | **66** | **0.3 (0.0 to 0.5)** | **0.042** | -0.2 (-0.6 to 0.2) | 0.27 | 60 | 0.0 (-1.0 to 0.9) | 0.97 | -0.2 (-1.7 to 1.3) | 0.80 | 38 | -0.2 (-1.5 to 1.1) | 0.79 | 1.2 (-0.3 to 2.8) | 0.12 |
|  | Sleep onset<sup>2</sup> | 45 | 0.1 (-0.1 to 0.4) | 0.34 | -0.2 (-0.8 to 0.3) | 0.43 | 42 | -0.3 (-1.2 to 0.6) | 0.53 | 0.0 (-1.9 to 2.0) | 0.95 | 26 | -0.8 (-2.2 to 0.7) | 0.25 | 0.4 (-2.3 to 3.1) | 0.72 |
|  | Sleep period | 66 | 0.3 (-0.1 to 0.6) | 0.12 | 0.1 (-0.2 to 0.5) | 0.42 | 60 | -0.4 (-1.8 to 1.0) | 0.57 | 1.1 (-0.3 to 2.5) | 0.11 | 38 | 0.1 (-1.3 to 1.5) | 0.89 | -0.4 (-2.2 to 1.4) | 0.68 |
|  | Sleep duration | 45 | 0.2 (-0.1 to 0.5) | 0.23 | 0.0 (-0.4 to 0.4) | 0.85 | 42 | 0.3 (-0.8 to 1.4) | 0.55 | 0.5 (-0.9 to 2.0) | 0.45 | 26 | -0.1 (-1.2 to 0.9) | 0.80 | -0.2 (-3.4 to 2.9) | 0.85 |

### White children

**Table 3** shows that for White children aged 36m there were no significant associations, although there was some indication that later sleep onset on weekends than weekdays was associated with smaller sum of 2-skinfolds. In children aged 18m, independent of all covariables including sleep period and duration, later daily sleep onset was associated with smaller sum of 2-skinfolds and lower BMI *z*-score (though further adjustment for final mealtime attenuated the latter association (−0.2 (−0.5 to 0.2), *p*=0.28)). Figure 1 illustrates that at age 18m, compared to children with a sleep onset time of 7:30pm or earlier, children who began to sleep between 7:30 and 8:30pm had smaller waist circumferences (−1.5 (−2.9 to −0.1) cm, *p*=0.035). Figure 2 highlights that children aged 18m who went to sleep ≥20 minutes later on weekends than weekdays, had smaller waist circumferences than children with consistent sleep onset times (−1.7 (−3.2 to −0.1) cm, *p*=0.038).

**Table 3.** Associations of sleep onset, period and duration with adiposity in White children, stratified by age group.

|  | Body mass index z-score |  |  |  |  | Waist circumference (cm) |  |  |  |  | Sum of 2-skinfolds (mm) |  |  |  |  |
| --- | --- | --- | --- | --- | --- | --- | --- | --- | --- | --- | --- | --- | --- | --- | --- |
|  | Weighted daily |  | average | Weekday to weekend difference |  | Weighted daily |  | average | Weekday to weekend difference |  | Weighted daily |  | average | Weekday to weekend difference |  |
|  | *n* | β (95% CI) | *p* | β (95% CI) | *p* | *n* | β (95% CI) | *p* | β (95% CI) | *p* | *n* | β (95% CI) | *p* | β (95% CI) | *p* |
| Model 1 |  |  |  |  |  |  |  |  |  |  |  |  |  |  |  |
| Sleep onset | 125 | -0.2 (-0.4 to 0.0) | 0.086 | 0.0 (-0.2 to 0.2) | 0.70 | 109 | -0.5 (-1.2 to 0.2) | 0.18 | -0.6 (-1.4 to 0.2) | 0.17 | 84 | -1.0 (-2.1 to 0.0) | 0.061 | -0.1 (-1.2 to 1.0) | 0.8 |
| Sleep period | 125 | 0.0 (-0.2 to 0.2) | 0.83 | 0.1 (-0.1 to 0.2) | 0.44 | 109 | 0.2 (-0.6 to 0.9) | 0.64 | 0.1 (-0.5 to 0.7) | 0.70 | 84 | 0.1 (-1.0 to 1.2) | 0.82 | -0.1 (-0.9 to 0.8) | 0.8 |
| Sleep duration | 108 | 0.0 (-0.2 to 0.2) | 0.91 | 0.1 (-0.1 to 0.3) | 0.37 | 93 | 0.1 (-0.7 to 1.0) | 0.74 | 0.3 (-0.4 to 1.1) | 0.39 | 76 | -0.1 (-1.0 to 0.8) | 0.82 | 0.1 (-0.9 to 1.1) | 0.8 |
| Model 2 |  |  |  |  |  |  |  |  |  |  |  |  |  |  |  |
| Sleep onset | 125 | -0.2 (-0.4 to 0.0) | 0.11 | 0.1 (-0.1 to 0.3) | 0.39 | 109 | -0.5 (-1.2 to 0.2) | 0.18 | -0.5 (-1.3 to 0.3) | 0.23 | 84 | -1.0 (-2.1 to 0.0) | 0.060 | 0.1 (-1.0 to 1.2) | 0.8 |
| Sleep period | 125 | 0.0 (-0.2 to 0.2) | 0.92 | 0.0 (-0.1 to 0.2) | 0.81 | 109 | 0.1 (-0.6 to 0.9) | 0.71 | 0.0 (-0.6 to 0.6) | 0.99 | 84 | 0.1 (-1.0 to 1.1) | 0.91 | -0.3 (-1.2 to 0.5) | 0.4 |
| Sleep duration | 108 | 0.0 (-0.2 to 0.2) | 0.99 | 0.0 (-0.1 to 0.2) | 0.62 | 93 | 0.2 (-0.6 to 1.1) | 0.57 | 0.2 (-0.6 to 1.1) | 0.55 | 76 | -0.1 (-1.0 to 0.8) | 0.86 | -0.2 (-1.3 to 0.9) | 0.7 |
| Model 3 |  |  |  |  |  |  |  |  |  |  |  |  |  |  |  |
| Sleep onset<sup>1</sup> | 125 | -0.2 (-0.5 to 0.0) | 0.072 | 0.2 (-0.1 to 0.5) | 0.15 | 109 | -0.6 (-1.5 to 0.3) | 0.18 | -0.7 (-1.7 to 0.3) | 0.15 | **84** | **-1.5 (-2.8 to -0.2)** | **0.025** | -0.2 (-1.6 to 1.2) | 0.7 |
| Sleep onset<sup>2</sup> | **108** | **-0.3 (-0.5 to -0.0)** | **0.044** | 0.0 (-0.2 to 0.3) | 0.79 | 93 | -0.6 (-1.6 to 0.3) | 0.20 | -0.5 (-1.4 to 0.4) | 0.28 | **76** | **-1.5 (-2.9 to 0.0)** | **0.046** | 0.0 (-1.3 to 1.2) | 0.9 |
| Sleep period | 125 | -0.1 (-0.3 to 0.1) | 0.41 | 0.1 (-0.1 to 0.3) | 0.31 | 109 | -0.2 (-1.5 to 0.3) | 0.69 | -0.3 (-1.0 to 0.4) | 0.40 | 84 | -0.8 (-2.1 to 0.5) | 0.22 | -0.6 (-1.6 to 0.4) | 0.2 |
| Sleep duration | 108 | -0.1 (-0.2 to 0.1) | 0.49 | 0.1 (-0.1 to 0.3) | 0.48 | 93 | 0.0 (-0.9 to 0.9) | 0.99 | 0.1 (-0.8 to 0.9) | 0.88 | 76 | -0.6 (-1.6 to 0.4) | 0.23 | -0.3 (-1.5 to 0.8) | 0.5 |
| Model 1 |  |  |  |  |  |  |  |  |  |  |  |  |  |  |  |
| Sleep onset | 96 | 0.0 (-0.2 to 0.3) | 0.71 | -0.1 (-0.3 to 0.1) | 0.32 | 96 | 0.2 (-0.7 to 1.1) | 0.63 | -0.1 (-0.9 to 0.8) | 0.89 | 67 | -0.3 (-1.6 to 0.9) | 0.57 | -0.6 (-1.6 to 0.5) | 0.2 |
| Sleep period | 96 | 0.0 (-0.3 to 0.3) | 0.98 | 0.0 (-0.2 to 0.2) | 0.91 | 96 | -0.5 (-1.6 to 0.5) | 0.31 | -0.3 (-1.1 to 0.5) | 0.40 | 67 | -0.2 (-1.6 to 1.2) | 0.82 | -0.1 (-1.1 to 0.9) | 0.8 |
| Sleep duration | 90 | -0.1 (-0.3 to 0.1) | 0.36 | 0.0 (-0.2 to 0.2) | 0.98 | 90 | -0.5 (-1.4 to 0.4) | 0.26 | -0.1 (-0.9 to 0.7) | 0.81 | 62 | -0.1 (-1.6 to 1.3) | 0.88 | 0.3 (-0.8 to 1.5) | 0.5 |
| Model 2 |  |  |  |  |  |  |  |  |  |  |  |  |  |  |  |
| Sleep onset | 96 | 0.0 (-0.2 to 0.3) | 0.87 | -0.1 (-0.3 to 0.1) | 0.38 | 96 | 0.1 (-0.9 to 1.0) | 0.90 | 0.0 (-1.0 to 0.9) | 0.92 | 67 | -0.3 (-1.6 to 1.0) | 0.68 | -0.9 (-2.1 to 0.2) | 0.1 |
| Sleep period | 96 | 0.1 (-0.2 to 0.4) | 0.66 | 0.0 (-0.2 to 0.2) | 0.81 | 96 | -0.4 (-1.5 to 0.8) | 0.51 | -0.2 (-1.1 to 0.6) | 0.54 | 67 | -0.1 (-1.6 to 1.3) | 0.86 | 0.1 (-1.0 to 1.1) | 0.8 |
| Sleep duration | 90 | -0.1 (-0.3 to 0.2) | 0.55 | 0.1 (-0.2 to 0.3) | 0.55 | 90 | -0.4 (-1.3 to 0.5) | 0.37 | 0.1 (-0.8 to 1.0) | 0.81 | 62 | -0.2 (-1.7 to 1.3) | 0.79 | 0.5 (-0.8 to 1.8) | 0.4 |
| Model 3 |  |  |  |  |  |  |  |  |  |  |  |  |  |  |  |
| Sleep onset<sup>1</sup> | 96 | 0.0 (-0.2 to 0.3) | 0.79 | -0.1 (-0.4 to 0.1) | 0.33 | 96 | 0.0 (-1.0 to 1.0) | 0.98 | -0.2 (-1.3 to 0.9) | 0.71 | 67 | -0.3 (-1.6 to 1.0) | 0.67 | -1.3 (-2.8 to 0.1) | 0.0 |
| Sleep onset<sup>2</sup> | 90 | 0.0 (-0.3 to 0.3) | 0.98 | 0.0 (-0.3 to 0.2) | 0.72 | 90 | 0.0 (-1.0 to 1.0) | 0.98 | 0.1 (-0.9 to 1.1) | 0.83 | 62 | -0.4 (-1.9 to 1.2) | 0.65 | -1.4 (-3.1 to 0.2) | 0.0 |
| Sleep period | 96 | 0.1 (-0.2 to 0.4) | 0.63 | 0.0 (-0.3 to 0.2) | 0.76 | 96 | -0.4 (-1.6 to 0.8) | 0.52 | -0.3 (-1.3 to 0.6) | 0.48 | 67 | -0.2 (-1.6 to 1.3) | 0.83 | -0.6 (-2.0 to 0.7) | 0.3 |
| Sleep duration | 90 | -0.1 (-0.3 to 0.2) | 0.55 | 0.0 (-0.2 to 0.3) | 0.68 | 90 | -0.4 (-1.4 to 0.5) | 0.37 | 0.1 (-0.8 to 1.1) | 0.76 | 62 | -0.3 (-1.8 to 1.3) | 0.74 | -0.4 (-2.0 to 1.3) | 0.6 |

### Sensitivity analyses

Further adjustment for physical activity in models that included children aged 36m, and height when waist circumference and the sum of 2-skinfolds were modelled as dependent variables, did not influence any of the reported associations. There was no evidence for interaction effects between sleep onset with the sleep period or sleep duration.

**Figure 1.**
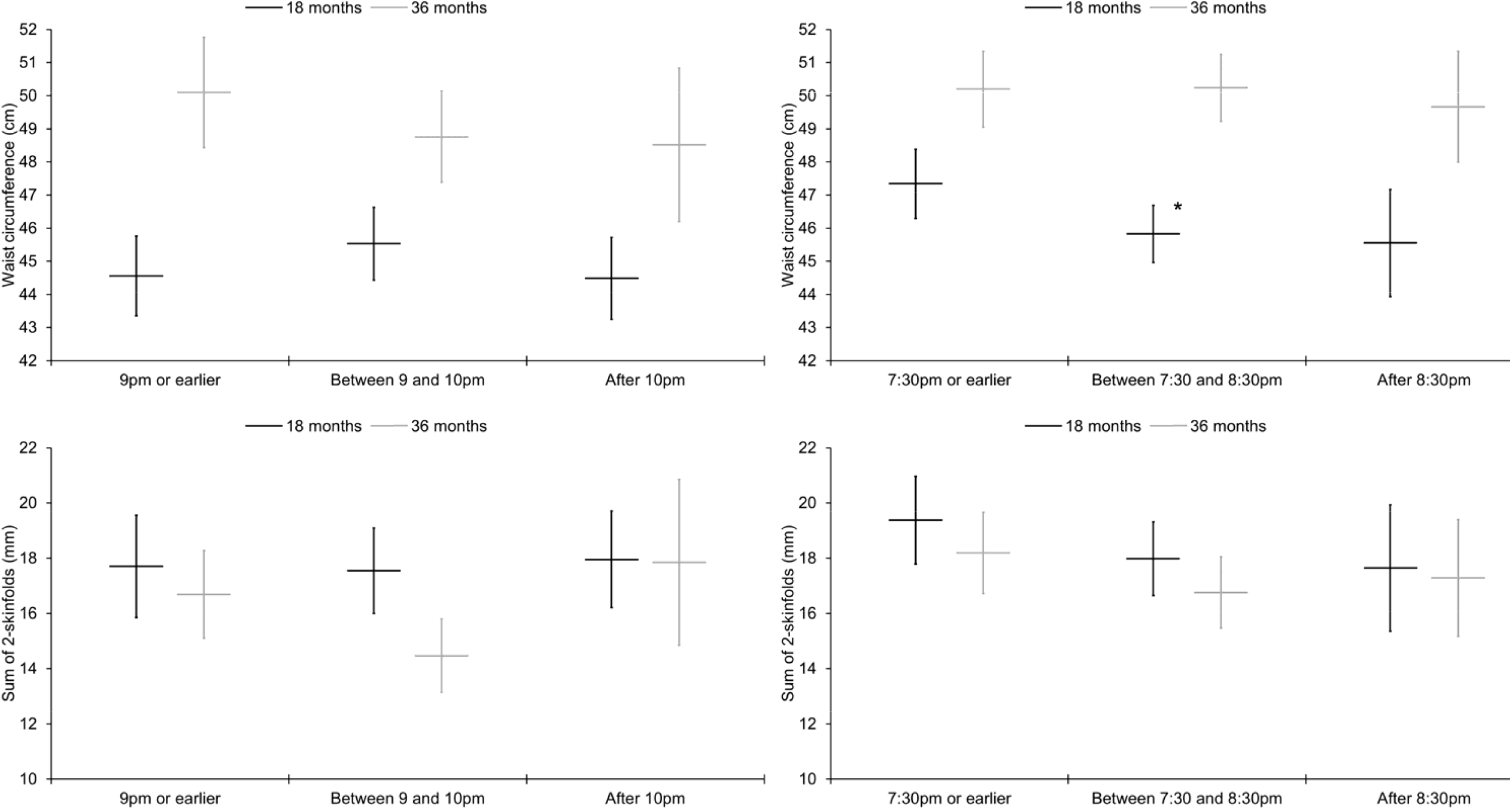
Associations of weighted daily average sleep onset categories with waist circumference and sum of 2-skinfolds, stratified by ethnicity and age group. Graphs on the left relate to South Asian children and graphs on the right to White children. Results are estimated marginal means (95% confidence intervals) adjusted for sex, age, socioeconomic status, maternal pregnancy age, parity, gestational age, birthweight, maternal BMI, season of measurement, napping frequency, unhealthy snacking, fruit and vegetable intake, TV viewing, and the sleep period. *Significantly different from the reference category of 7:30pm or earlier (β=-1.5 (−2.9 to −0.1) cm, *p*=0.035); significant difference persisted when further adjusted for final mealtime (β=-1.7 (−3.2 to −0.1) cm, *p*=0.036). Results are presented for South Asian children aged 18m (9pm or earlier: waist circumference, *n*=25; skinfolds, *n*=19; Between 9 and 10pm: waist circumference, *n*=26; skinfolds, *n*=23; After 10pm: waist circumference, *n*=26; skinfolds, *n*=22) and 36m (9pm or earlier: waist circumference, *n*=22; skinfolds, *n*=16; Between 9 and 10pm: waist circumference, *n*=28; skinfolds, *n*=17; After 10pm: waist circumference, *n*=10; skinfolds, *n*=5), and White children aged 18m (7:30pm or earlier: waist circumference, *n*=39; skinfolds, *n*=30; Between 7:30 and 8:30pm: waist circumference, *n*=51; skinfolds, *n*=37; After 8:30pm: waist circumference, *n*=19; skinfolds, *n*=17) and 36m (7:30pm or earlier: waist circumference, *n*=36; skinfolds, *n*=25; Between 7:30 and 8:30pm: waist circumference, *n*=42; skinfolds, *n*=29; After 8:30pm: waist circumference, *n*=18; skinfolds, *n*=13).

**Figure 2.**
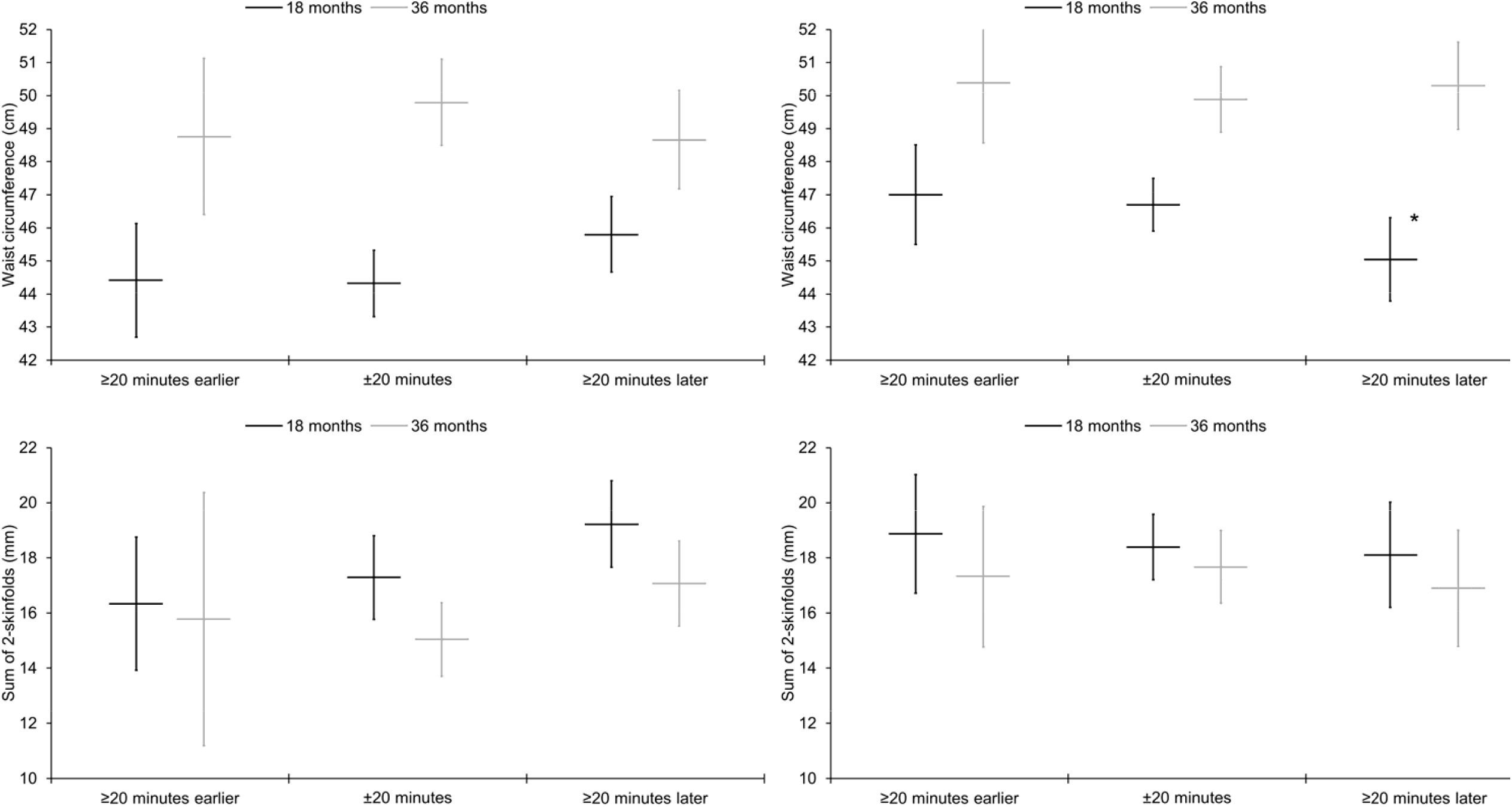
Associations of weekday to weekend differences in sleep onset with waist circumference and sum of 2-skinfolds, stratified by ethnicity and age group. Graphs on the left relate to South Asian children and graphs on the right to White children. Results are estimated marginal means (95% confidence intervals) adjusted for sex, age, socioeconomic status, maternal pregnancy age, parity, gestational age, birthweight, maternal BMI, season of measurement, napping frequency, average weekday sleep onset, unhealthy snacking, fruit and vegetable intake, average weekday TV viewing, week to weekend differences in TV viewing, average weekday sleep period, and week to weekend differences in the sleep period. *Significantly different from the reference category of ±20 minutes (β=-1.7 (−3.2 to −0.1) cm, *p*=0.038). Results are presented for South Asian children aged 18m (≥20 minutes earlier: waist circumference, *n*=16; skinfolds, *n*=15; ±20 minutes: waist circumference, *n*=34; skinfolds, *n*=27; ≥20 minutes later: waist circumference, *n*=27; skinfolds, *n*=22) and 36m (≥20 minutes earlier: waist circumference, *n*=10; skinfolds, *n*=3; ±20 minutes: waist circumference, *n*=28; skinfolds, *n*=21; ≥20 minutes later: waist circumference, *n*=22; skinfolds, *n*=14), and White children aged 18m (≥20 minutes earlier: waist circumference, *n*=20; skinfolds, *n*=16; ±20 minutes: waist circumference, *n*=60; skinfolds, *n*=46; ≥20 minutes later: waist circumference, *n*=29; skinfolds, *n*=22) and 36m (≥20 minutes earlier: waist circumference, *n*=19; skinfolds, *n*=14; ±20 minutes: waist circumference, *n*=47; skinfolds, *n*=35; ≥20 minutes later: waist circumference, *n*=30; skinfolds, *n*=18).

## Discussion

This study investigated associations of diarised sleep onset time, period and duration with total and central adiposity in young children aged 18 and 36m. We discovered ethnicity- and age-specific associations for sleep onset times, which we primarily attribute to markedly different sleep schedules between South Asian and White children. Later daily sleep onset, and later sleep onset on weekends than weekdays, were both associated with higher adiposity in South Asian children and conversely lower adiposity in White children. Longer sleep duration on weekends than weekdays was associated with higher adiposity in South Asian children aged 18m.

Numerous systematic reviews have summarised the evidence for associations of sleep duration or the sleep period with adiposity in children and adolescents [1,2]. Of note, one review meta-analysed the results of six prospective studies totalling 14,264 children aged <36m. There was moderate heterogeneity in results, but each additional hour of sleep was associated with a negative change in BMI z-score over follow-up [36]. Limitations of the existing evidence include a preponderance of studies that have relied solely on BMI or BMI z-score as proxies for total adiposity. Studies have also considered few potential confounding or mediating factors and focussed solely on sleep duration. Sleep is a multidimensional construct and it is important to unravel the independent relations of specific dimensions with adiposity. In a previous BiB 1000 study we analysed repeated questionnaire data collected at four timepoints (12, 18, 24 and 36m) to quantify associations between parent reported sleep duration with measured adiposity in 1338 UK South Asian and White children. Longer sleep duration predicted lower total and central adiposity, but only in South Asian children [20]. Here, in a sub-sample of the same cohort, we were able to scrutinise and mutually adjust for myriad sleep parameters collected over the course of three days via parent-completed sleep diaries. Independent of sleep length, the average sleep onset time and variability in sleep timing between weekdays and weekends emerged as the key modifiable dimensions that predicted adiposity in South Asian and White children.

A recent review concluded that sleep timing, in particular later bedtimes, is associated with higher weight status in primary school-aged children [9]. Similarly, the few studies conducted thus far in 4 to 5 year olds indicate that later bedtimes, and most markedly bedtimes after 9pm, are associated with higher BMI z-score and obesity risk [5–7]; one study found that an association of short sleep with higher BMI z-score was only evident in children who went to bed after 9pm [8]. Just one study to date has been performed in younger children. Roy et al. found that later bedtimes were associated with higher odds of obesity in children aged 36m, but the association attenuated when adjusted for sleep duration [17]. To our knowledge, the current study is the first to show that independent of the sleep period, later sleep onset was associated with higher BMI z-score in South Asian children aged 36m. The association attenuated when adjusted for final mealtime, which provides some indication that later sleep onset may elevate obesity risk via later evening meals and higher caloric intake later in the day [37]. In line with this hypothesis, animal models and clinical trials have shown that early and time-restricted feeding has numerous physiological benefits, including lower adiposity [38,39].

With regard to weekday and weekend differences, a recent review and pooled analysis of five studies in school-aged children and youth indicated that later sleep timing on weekends than weekdays was correlated with higher risk of overweight or obesity, albeit with a small effect size [10]. Our results extend this observation to younger children of South Asian heritage. We uniquely observed that later sleep timing on weekends than weekdays was associated with higher adiposity in South Asian children aged 18m and larger sum of 2-skinfolds in South Asian children aged 36m, although the association in older children only approached statistical significance. The associations in children aged 18m were independent of potential confounding or mediating factors including TV viewing, unhealthy snacking, fruit and vegetable intake, and the final mealtime. It may be that these are not important mediators in very young children, who are less autonomous, and for whom a large part of their diet and screen time is governed by parents and carers. That said, we cannot exclude residual confounding or mediation effects, as the data were all parent-reported and are subject to random error and bias. It is also unfortunate that physical activity was not measured at 18m. Longer sleep duration on weekends than weekdays was associated with higher adiposity at South Asian children aged 18m. This suggests that consistent sleep durations are important.

We observed that later daily sleep onset was associated with lower BMI z-score in White children aged 18m. This observation may appear counterintuitive, but Roy et al. similarly reported that independent of sleep duration, each one hour later to bed was associated with-0.08 lower BMI z-score in 878 children, although their confidence interval did narrowly cross the null of no association (95% CI: −0.17 to 0.01) [17]. Following adjustment for final mealtime our result for BMI z-score also attenuated to the null, but inverse associations with more direct adiposity indicators remained, including smaller sum of 2-skinfolds and waist circumference as a function of later sleep onset. These unexpected results may partly be explained by early sleep onset obstructing time during early-to-mid evenings that could otherwise be spent physically active. Time-stamped accelerometer studies show that daily physical activity patterns are bimodal in 18m olds. Physical activity levels first peak in the morning, slump after midday due to feeding and napping (81% of our White children aged 18m napped every day), after which there is a second larger peak that begins around 4pm and tails-off by 8pm [40,41]. In our sample of White children aged 18m, the earliest sleep onset times were around 6pm and more than one-third (36.8%) were asleep by 7:30pm. It is conceivable that these children may have missed a substantial part of the evening peak in physical activity, thus contributing to higher adiposity [29,42]. Our related observation in the same group of children, that going to sleep ≥20 minutes later on weekends than weekdays was associated with lower waist circumference, may be explained by later sleep onset at the weekend enabling more physical activity [43].

Although White children aged 36m exhibited a similar distribution of sleep onset times compared to their younger White peers, there were few comparable associations, except some indication in children aged 36m that later sleep onset on weekends than weekdays was associated with lower adiposity (an association with the sum of 2-skinfolds approached statistical significance). These age-related differences may be explained by daily physical activity patterns following a different trajectory in slightly older children who tend not to nap (just 5% of White children aged 36m napped every day; 46% napped occasionally; 49% never napped). Instead of a naptime-related slump after midday which is followed by an evening peak, physical activity levels tend to be relatively constant throughout the afternoon, before progressively declining in the evening and most notably from 7pm onwards [44]. Hence, in many White children aged 36m, there may have been no peak in evening physical activity for early sleep to obstruct. In contrast to our results, Roy et al. did find a significant association between later bedtimes with lower BMI z-score in children aged approximately 36m [17]. That study was performed in children living in America, Australia and New Zealand, which likely explains the discrepancy. In those countries frequent daytime naps persist in older children partly because of designated naptimes in childcare [45–47], hence many children aged 3 to 4 years continue to exhibit a bimodal physical activity pattern, including an early-to-mid evening activity peak that early sleep could obstruct [48].

Sleep recommendations for children have historically focussed on duration rather than other sleep dimensions [49]. Our results highlight that contemporary sleep guidelines should further acknowledge the importance of sleep timing and regular sleep schedules [50]. Any such recommendations should also be considerate of ethnic and cultural differences in sleeping patterns, including what may and may not be feasible and acceptable to different population subgroups. Our data highlight that parents of White children should be made aware that going to sleep too early may be detrimental, certainly from an energy-balance and obesity risk perspective, and that going to sleep before 7:30pm might be inadvisable. Parents of South Asian children could be encouraged to set an earlier evening meal, an earlier sleep onset time, and to uphold consistent sleep schedules across week and weekend days. Studies of 4 to 5 year olds indicate that bedtimes before 9pm are important [5–8]. Our data also suggest that a reasonable and achievable target for young South Asian children might be a sleep onset time before 9pm. It could be argued that any such changes in sleeping patterns may have only a trivial impact on adiposity, because the associations we have reported are modest in scale. However, our associations might be underestimated due to measurement imprecision and daily fluctuations in sleeping patterns. Furthermore, any influence on adiposity levels in early childhood may be important, it is a critical period in which rapid weight gain occurs and sustained obesity develops [16].

This study uniquely investigated associations of diarised sleep parameters with myriad adiposity markers in a biethnic sample of young children from a deprived urban setting, a high-risk group for sleep problems and obesity [18,19]. Sleep diaries are superior to the common practice of posing a single question to parents about their child’s ‘usual’ sleep [51]. They do however incur participant burden and it is unfortunate that a minority of BiB 1000 parents completed sleep diaries for their child, the consequences of which are at least twofold. First, it is probable that our study sample is subject to selection biases which hinders generalisability. Second, stratifying by ethnicity and age group meant that some of our analyses were based on relatively few observations and low statistical power. This latter issue was compounded by missing data for specific items, such as sleep duration (which was only reported if parents considered they knew how long their child had slept overnight) and the sum of 2-skinfolds. It is nevertheless advantageous that we investigated more direct adiposity markers than suboptimal weight-for-height proxies and that in addition to skinfolds we investigated waist circumferences; young South Asian children eventually develop more centrally-stored adiposity which is metabolically deleterious [22]. Another strength is that each of our analyses were adjusted for a broad range of covariates, including mutual adjustment for sleep parameters to tease apart independent associations of distinct sleep dimensions with adiposity. Unfortunately, directions of association cannot be inferred from this cross-sectional study, it is possible that adiposity could influence sleeping habits rather than vice versa [20]. Longitudinal studies or trials are needed to determine the direction of associations. Future studies may achieve a better trade-off between participant retention and data resolution if they were to use sensor-based technologies rather than diaries [52], alternatively a combination of methods may prove best to capture habitual sleep parameters [53].

## Conclusions

Sleep onset time appears to be independently associated with levels of total and central adiposity in young South Asian and White children. Suitable sleep times and consistent sleep schedules across week and weekend days could be important modifiable determinants of early onset obesity.

## Data Availability

Data generated and analysed for the current study are available from the corresponding author on reasonable request.

## Notes

### Competing Interest Statement

The authors have declared no competing interest.

### Funding Statement

The Born in Bradford study receives core infrastructure funding from the Wellcome Trust (grant number: WT101597MA), a joint grant from the UK Medical Research Council (MRC) and UK Economic and Social Science Research Council (ESRC) (MR/N024397/1), the British Heart Foundation (BHF) (CS/16/4/32482), and the National Institute for Health Research (NIHR) under its Collaboration for Applied Health Research and Care (CLAHRC) for Yorkshire and Humber. This study received delivery support from the NIHR Clinical Research Network. John Wright leads the Healthy Children, Healthy Families Theme of the NIHR CLAHRC in Yorkshire and Humber. Paul J Collings is funded by a BHF Immediate Postdoctoral Basic Science Research Fellowship (FS/17/37/32937). Jane E Blackwell coordinates and Paul J Collings is a member of The White Rose Child & Adolescent Sleep Research Network which is funded by a White Rose Collaboration Grant. The views expressed in this paper are those of the authors and not necessarily those of the MRC, ESRC, BHF, NIHR, and UK Department of Health or National Health Services or of any other funder acknowledged here.

### Author Declarations

Ethical approval for all aspects of the study was granted by Bradford Research Ethics Committee (Reference 07/H1302/112).

